# Machine learning-enhanced behavioural approach to detecting reactions to sound in infants and toddlers: proof-of-concept study

**DOI:** 10.1101/2025.07.10.25331271

**Authors:** Chelsea M. Blankenship, Josef Schlittenlacher, Iain R. Jackson, Anisa S. Visram, Kevin J. Munro, Lisa L. Hunter, David R. Moore

## Abstract

**Objective:** Show that a basic unsupervised machine learning (ML) algorithm can give information on the direction of child and infant reactions to sound using non-identifiable video-recorded facial features.

**Design:** Infants and toddlers were presented warble tones or single-syllable utterances 45 degrees to the left or right. A camera recorded their reactions, from which features like head turns or eye gaze were extracted with OpenFace. Three clusters were formed using Expectation Maximization on 80% of the toddler data. The remaining 20% and all infant data were used to verify if the clusters represent groups for sound presentations to the left, to the right, and both directions.

**Study Sample:** 28 infants (2-5 months) and 30 toddlers (2-4 years), born preterm (<32 weeks gestational age) were presented ten sounds each.

**Results:** The largest cluster comprised 90% of the trials with sound presentations in both directions, indicating “no decision.” The remaining two clusters could be interpreted to represent reactions to the left and the right, respectively, and average sensitivities of 96% for the toddlers and 68% for the infants.

**Conclusions:** A simple machine learning algorithm demonstrated that it can form correct decisions on the direction of sound presentation using non-identifiable facial behavioural data.

## 1. Introduction

Congenital hearing loss is among the most common conditions at birth that result in adverse neurodevelopmental outcomes, including language, academic, and social skill deficits (Moeller et al., 2007; Yoshinaga-Itano et al., 2021). Infants at highest risk for hearing loss are those born very prematurely who are cared for in neonatal intensive care units (NICU) (Beswick et al., 2012; Hirvonen et al., 2018; Hunter et al., 2023). Fortunately, congenital hearing loss is effectively addressed through universal newborn hearing screening programs, if diagnosis and intervention are provided within the first few months after birth (Ching et al., 2017; Sininger et al., 2009; Tomblin et al., 2015; Yoshinaga-Itano et al., 1998).

### 1.1 Paediatric Behavioural Hearing Assessment Methods

Behavioural assessment of infants’ responses to sound is crucial to validate electrophysiologic hearing assessments and determine the need for, and effectiveness of, auditory intervention. Based on a large study of developmental milestones, infants alert to sound by the age of 1 week, and orient towards sounds by 4 months (Capute et al., 1986). Behavioural observation of subtle infant responses to sounds have long been used for behavioural observation audiometry (BOA) (Bench et al., 1976a, 1976b, 1976c). BOA is an unconditioned procedure that relies on behavioural response from the infant, such as changes in sucking, eye shifts, forehead wrinkling or head orientation, in response to sound. However, this behavioural observation method requires a highly trained observer, and success is dependent on the child’s age, developmental level, attention span, and cooperation (Widen & Keener, 2003). BOA has high inter- and intrasubject variability in responses and rapid response habituation (Hicks et al., 2000) and it is difficult to control for observer bias in a clinical setting (Widen, 1993). Therefore responses obtained with BOA are not considered suitable or reliable enough for clinical use (Ruth et al., 1983).

For research purposes, an observer-based psychoacoustic procedure (OPP) was developed that requires highly trained human observers and specialized software (Olsho et al., 1987; Werner & Marean, 1991). OPP controls observer bias by providing auditory masking of the observer, and a two-alternative forced choice response paradigm with psychoacoustic tracking, allowing testing in very young infants that are not yet developmentally ready for VRA testing (Olsho et al., 1988). However, despite the use of trained observers, many very young infants do not respond with adequate reliability, and they have poorer thresholds than children and adults, either due to inattention or different listening strategies. Thus, OPP is generally unsuitable for clinical application.

From around a developmental age of 6-7 months, infants start to reliably respond to sound input with reinforced head turns. At this age, visual reinforcement audiometry (VRA) is commonly used to diagnose hearing loss and track intervention outcomes (Widen & Keener, 2003). VRA involves conditioning the child to make a head turn response following the presentation of an auditory stimulus (detection not localization task). Most commonly, this involves the infant looking towards a visual target such as a toy or video monitor, but immature or subtle responses can also include eye shifts/movements and stilling or quieting. VRA is generally recommended for infants and young children between 6 months to 2.5 years of age, since some older children may lose interest in the reinforcement or are too active and need a task to maintain attention. Despite its widespread use, VRA has limitations: responses can be influenced by the child’s developmental level, fatigue, or distractibility. Additionally, false positives may occur if the child anticipates reinforcement rather than responding to the auditory stimulus, and testing can be challenging in children with motor delays, limited visual attention, or visual impairments. Lastly, VRA typically involves two trained individuals making it more resource-intensive which could present as a logistical challenge in busy clinical settings or small clinics with limited personnel.

Conditioned Play Audiometry (CPA) is a behavioural hearing test typically used for children between approximately 2.5 and 5 years of age, when they are developmentally ready to engage in structured play tasks. CPA involves conditioning the child to perform simple task like stacking rings, building towers, etc., every time they hear an auditory stimulus. Transforming the hearing test into a game improves attention and motivation resulting in more reliable threshold information across a wider range of test frequencies compared to VRA (Thompson & Weber, 1974; Thompson & Thompson, 1972). However, successful CPA requires that the child understands the task, has the control to wait for the stimulus before responding, which can be challenging for children with developmental delays, attention difficulties, or limited motor skills.

In summary, while there are several behavioural methods that are used clinically to assess hearing in infants and young children, they each have limitations and challenges, and success is highly dependent on the child’s temperament, attention, and ability to engage with the task. Additionally, many children with hearing loss have comorbid conditions such as developmental delays or neurocognitive disorders, which make it difficult to complete behavioural assessments. Consequently, assessing hearing in very young infants and in difficult-to-test populations remains a significant clinical challenge. To address these gaps and improve diagnostic accuracy, it is essential to explore objective approaches for hearing assessments that don’t rely on observers’ subjective interpretation of behavioural responses.

### 1.2 Artificial Intelligence and Auditory Research

The use of artificial intelligence (AI) in medical settings is becoming increasingly common and is rapidly transforming healthcare delivery and medical research. AI is a broad term that encompasses several subsets, including machine learning (ML; system learns from data and makes predictions without explicit programming), deep learning (a subset of ML that uses multi-layered artificial neural networks) and, more recently, generative AI (develops new content such as text, audio, pictures., etc.) and large language models. Furthermore, it’s well established that AI has the potential to improve medical care by improving diagnostic accuracy, increasing treatment efficiency, and ultimately optimizing patient outcomes (Chinta et al., 2025).

Despite the advantages of AI, its use in healthcare and research presents inherent challenges related. First, many ML techniques require large datasets—often tens of thousands of measures for basic algorithms and millions for complex image recognition tasks (Deng et al., 2009; Krizhevsky, 2009). In paediatric hearing research, such datasets are unavailable and would require extensive manual labelling by human observers. Second, ethical considerations around data privacy are significant. High-performing, end-to-end deep learning models typically rely on raw images, which cannot be fully anonymized because facial features and other identifiers remain embedded in the data. This raises concerns about compliance with privacy regulations and participant confidentiality. To mitigate these risks, studies should prioritize storing only low-dimensional, non-identifiable representations of responses rather than raw images.

OpenFace (Baltrusaitis et al., 2019; Baltrušaitis et al., 2015) is an open-source software toolkit for analysing video data. It can be used to derive standardised behavioural features from faces detected in video recordings, including three-dimensional head position, head rotation across three different axes, eye gaze direction, and activation of facial action units (e.g., eyebrow raising). In contrast to facial landmarks that encode where specific parts of the face are located, these variables are less likely to allow identification of a child. Additional processes, such as reducing the data with a principal component analysis (PCA), can further ensure privacy. Future work should focus on approaches that use a limited set of anonymized behavioural indicators, while building justification for collecting large, clinically relevant datasets that could later support more sophisticated ML solutions, including for example deep neural networks, for clinical implementation.

Within the field of audiology, there has been a recent influx of publications using AI approaches (Frosolini et al., 2024). In a systematic review of AI in audiology, Frosolini and colleagues identified 104 publications since 1990, with the majority (87.5%) published after 2020. Most publications focused on therapeutic and prognostic applications (34.6%), hearing assessment (33.7%), unique applications and exploratory research (21.1%), temporal bone radiology (5.8%) and vestibular diagnosis (5.8%). With regard to hearing assessments, manuscripts were published on a variety of topics ranging from hearing screening and diagnostic assessment (Gathman et al., 2023; Liu et al., 2021; Xie et al., 2025), measurement of middle ear function and diagnosis of otitis media (Peng et al., 2025; Zeng et al., 2022), hearing preservation (Zeitler et al., 2024) and post-implantation programming (Wathour, Govaerts, Derue, et al., 2023; Wathour, Govaerts, Lacroix, et al., 2023). However, most of the auditory AI research focusses on adults and older children. Infant AI literature primarily focuses on classifying middle ear function and analysis of electrophysiologic assessments including interpretation of auditory brainstem responses (McKearney et al., 2022), categorisation of neural responses in infants to drumbeats versus repeated syllables (Gibbon et al., 2021) and to pitch processing in neonates (Hart & Jeng, 2021). Currently, there are few publications using AI approaches to detect behavioural responses to sound in young children (Xie et al., 2025), none of which are in infants. Behavioural hearing assessments in this population have inherent challenges, creating a critical need for alternative approaches. Leveraging AI and machine learning (ML) may offer a solution for objectively measuring sound detection in infants. If successful, it has the potential to improve infant assessment methods which would ultimately impact diagnosis, guide treatment and improve outcomes.

### 1.3 Study Purpose and Research Questions

This paper presents an overview of our initial efforts to measure sound detection in infants and toddlers using ML to identify eye, facial and body movements in response to tonal and speech sounds. In this approach, a video camera replaces the human observer, and an algorithm trained on validated data determines whether observed movements indicate a reaction to sound. Specifically, we examine optical scanning of the infant’s face and torso combined with ML to monitor responses to sounds, including subtle cues a human observer might not reliably detect. Therefore, the overall purpose of this study was to investigate whether a behavioural observation-like auditory task could be trained with ML methods to learn when an infant shifted eye gaze, turned their head, or reacted with facial expressions, such as eye widening or eyebrow raising, in response to sounds. The study included a cohort of infants (2-5 months) and toddlers (2-4 years) who were born preterm and underwent hearing assessments on the same day as the ML protocol. In this proof-of-concept study, the primary research questions were: (1) can ML be used to cluster responses based on head and facial changes of infants and toddlers to presented sound stimuli? (2) can the trained algorithm accurately detect the direction of sound presentation when infants and toddlers respond to sound?

## 2. Method

### 2.1. Study Design and Participants

Participants were infants and toddlers born very or extremely preterm (< 32 weeks gestational age or less) recruited soon after birth from NICUs in Cincinnati, Ohio, United States and were enrolled in a longitudinal project to improve early detection of speech and language delays. This population is of research interest due to their higher risk for hearing loss and known difficulties with behavioural assessment due to more prevalent developmental delay. Infants with known chromosomal abnormalities, or congenital conditions affecting the central nervous system were excluded. All families were primarily English-speaking and agreed to have their child video and audio recorded.

Participants included 58 children split into an infant group (n = 28, mean = 3.0 mos., range = 2.1 - 4.6 mos.) and a toddler group (n = 30, mean = 29.8 mos., range = 23.9 - 41.7 mos.). All ages are reported corrected for gestational age. All participants were seen once for the ML study in conjunction with auditory brainstem response testing (infants) or age-appropriate behavioural audiometry (toddlers). Infants had normal hearing in both ears as determined with 1 kHz chirp-evoked auditory brainstem response thresholds (≤ 20 dB eHL)(Sininger et al., 2020). For toddlers, hearing status was based on VRA or conditioned play audiometry (CPA) thresholds, using earphones or sound field speakers with live speech and pure tone stimuli (1, 2, 4, and 8 kHz)(Hunter et al., 2023). If only sound field testing was completed, ear specific hearing status was determined through a combination of 226-Hz tympanometry, distortion product otoacoustic emissions (2-10 kHz) measured at the same visit along with repeated behavioural hearing assessment at follow-up study visits. Toddlers had normal hearing in both ears (n=29, ≤ 20 dB HL), except one toddler with mild conductive hearing loss (thresholds at 30 to 35 dB HL). The study was approved by the Institutional Review Board of Cincinnati Children’s Hospital Medical Center. Informed consent was obtained from the caregiver of all children prior to enrolment.

### 2.2. Apparatus

All testing was completed in a sound-treated booth using a custom Python program. The auditory stimuli were amplified (Crown D-75A, Harman) and presented through one of two sound field speakers (Control 1Xtreme, JBL). Sound field speakers were positioned at 45 degrees to the left and right at ear level at approximately 1 meter from the infant/toddler. A computer monitor was positioned directly in front of the child, at the midpoint between the two sound field speakers. To record the child’s responses, a video camera (AF/FHD1080P, FUVISION) was positioned underneath the monitor (see Figure 1).

**Figure 1.**
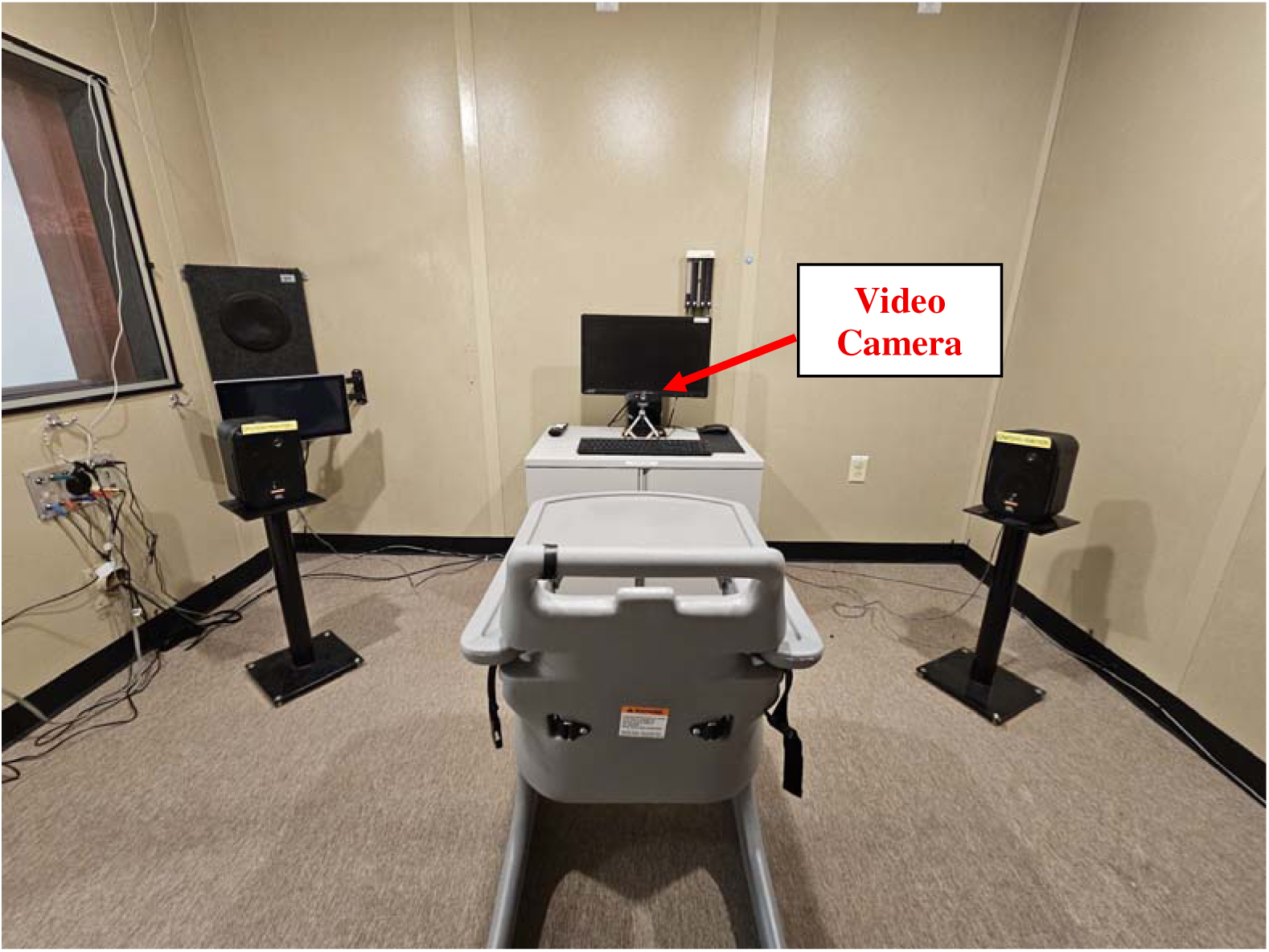
Experimental setup for toddlers who were seated in the high chair with sound field speakers positioned to the right and left at 45° azimuth relative to the ears at the same height. A computer monitor was straight ahead and played a centring video continuously throughout the test. The video camera was positioned underneath the computer monitor.

### 2.3. Stimuli

The children were presented five different auditory stimuli including frequency-modulated warble tones (duration of 500 msec, 5 Hz modulation rate, 5 % modulation depth, carrier frequencies: 0.5, 1, 2 kHz), and utterances (“ba” and “da”) through the right or left sound field speakers at a level of 60 dB SPL, approximating conversational speech levels. Each stimulus was presented twice in a randomized order, from either the left or right speaker. The stimuli were calibrated using a Larson Davis System 824 sound level meter (Depew, New York) and Brüel & Kjær ½” microphone.

### 2.4. Procedure

Toddlers sat in a highchair or, if restless, on their caregiver’s lap while infants were always positioned in their caregiver’s lap (see Fig. 1). If the caregiver was in the video recording space, they wore a face mask covering their nose and mouth to ensure the analysis software did not detect and include their face in the collected data. Each child was tested in a single session that started with an auditory visual primer: a black-and-white smiling face displayed on the computer monitor while the phrase “Hello Baby” was played through both sound field speakers to centre the child’s gaze forward. For infants, a slowly rotating shape with primary colours was presented throughout the test session to help maintain the infants’ attention. For the toddlers, a slowly moving colourful scene was presented throughout after the auditory primer. The test assistant (seated inside the booth next to the computer monitor) re-centred the child if needed between stimuli, ensured the child was upright, and could see the computer monitor. When the child was centred and quiet, the test assistant manually triggered the presentation of each sound (speech or tone) using a keypress. Responses to sounds were not rewarded, as would be done in a typical clinical reinforced procedure. This simplified approach was used to determine whether ML could detect spontaneous infant and toddler responses to sound, rather than trained responses and reinforced head turns. This was necessary for the proof-of-concept that naturalistic responses other than head turns could be detected. The inter-stimulus interval was set to a minimum of 6 seconds. In some cases, a longer interval was needed to re-centre the child between presentations. In this protocol, test sessions lasted 114 seconds on average (SD = 27 sec; range = 81 - 287). All participants included in the analysis successfully completed the test protocol in one session.

### 2.5. Data preprocessing with OpenFace

To protect participant privacy and avoid sharing raw images or videos for machine learning analysis, all video data were stored internally. Each video image was processed using OpenFace (Baltrušaitis et al., 2019), resulting in 55 standardised features per frame of video. These features included estimates on the degree of activation of 35 facial action units (Baltrušaitis et al., 2015), for example whether a part of an eyebrow is raised and by how much. The remaining features encoded the gaze (8 features: x, y, and z directions for each eye separately, and x and y angles each averaged across the two eyes (Wood et al., 2015), the location and rotation of the head (6 features: 3 coordinates and 3 angles, one for each spatial dimension/axis) and 6 features for the head shape. After an analysis of the reaction time to stimulus onsets (see section Results), these 55 features were stored for a single frame of the video, 2 seconds after stimulus onset. For this dataset, a maximum of 580 cases were obtained (58 participants × 10 stimulus presentations per participant). However, for 20 cases no data were available for 2 seconds after stimulus onset (2 infant cases, 18 toddler cases; i.e., no behavioural response following sound presentation). Therefore, a total of 560 cases or sets of features (278 infant cases, 282 toddler cases) were included in the analysis.

### 2.6. Clustering with Expectation Maximization (EM)

The need for annotation of the data for the training of an algorithm was avoided by using unsupervised ML and verifying it via the direction of sound presentations.

Unsupervised clustering ML algorithms learn structures in the data, without the need for labels, and allocate each measure to a cluster. The algorithm was trained to recognize three clusters with facial features as the defining measure. The meaning of the clusters thus formed was verified by matching the three predicted clusters to reactions to sound presentations from the left, the right, or no decision for either direction.

Expectation maximization (EM, (Dempster et al., 1977) was used to model the data by multivariate Gaussian distributions. Thus, each cluster is represented by its mean and spread. EM not only learns the spread of the data in each dimension (e.g., head turn and eye gaze across the vertical axis), but also covariances between these dimensions. For example, turning the head to the right/left is likely to be correlated with a gaze to the right/left, and thus would not give much additional information. EM updates its belief about the parameters iteratively and learns on its own to match these distributions to the collected data. In brief, it starts with initial means and covariances for each cluster and then assigns all data points to a cluster (the expectation step). Thereafter, it updates the means and covariances of each cluster based on the data that are presently predicted to belong to that cluster (the maximization step). These two steps are repeated until the parameters converge.

The algorithm was trained on 80% (n=226) of the participants from the toddler group. The remaining 20% (n=56) of the toddler group and all observations of the infant group (n=278) were used as validation sets. The reason for using only toddler data for training was because we expected more reliable responses from them. In particular, the infants might only be able to perform a subset of the actions that toddlers can or not turn their heads at all.

EM was applied to four different sets of two features each: (1) Rotation of the head along the vertical axis (yaw, turning left or right) and x-axis (pitch, nodding), (2) Gaze direction in world coordinates of the left eye to the left/right and up/down, (3) Gaze angle averaged across both eyes to the left/right and up/down, (4) the first two components of a principal component analysis of all 55 features. For analyses 1 to 3, it could be expected that the first variable that encodes looking left and right is most descriptive on reactions to the sound presentations from the left or right, but the second variable could potentially give general information about a reaction in any direction as well.

EM was performed using the R EMCluster package (Chen & Maitra, 2015). Since the results of the EM algorithm depend on a random initialization, the following procedure was followed to ensure replicable and consistent results: First, the algorithm was performed five times, and the best performing solution was chosen (using the function “exhaust.EM”) for the initialization of an EM using the function “shortemcluster” and a convergent tolerance of 0.0001. Except for the analysis on the PCA, this was repeated 100 times or until the maximum of all mean coordinates was at least half as large as the minimum and of opposite sign, and vice versa. This was done to encourage the algorithm to have two clusters that are of opposite sign, even though we did not enforce this to be in a particular or the same dimension. After this, the function “emcluster” was run with default parameters to determine the three clusters.

## 3. Results

### 3.1. Reaction Time and Absolute Head Angle Analysis

To determine if and when the children reacted to stimuli, the absolute angle of the head looking left or right was analysed as a function of time after stimulus onset. Figure 2 shows that immediately before stimulus onset, both toddler and infant head angle was about 10 degrees off-centre on average (mean of absolute values). The toddler group showed a maximum reaction 2 to 3 sec after stimulus onset, with an average head turn of 17 degrees (0.30 radians), while the infant group only slightly or rarely turned their head, to about 12 degrees (0.21 rad) on average, also at around 2 seconds. In light of these results, we decided to analyse the data from a single frame 2 seconds after stimulus onset and to use the toddler group, with their stronger head reactions, for training the EM algorithm.

**Figure 2.**
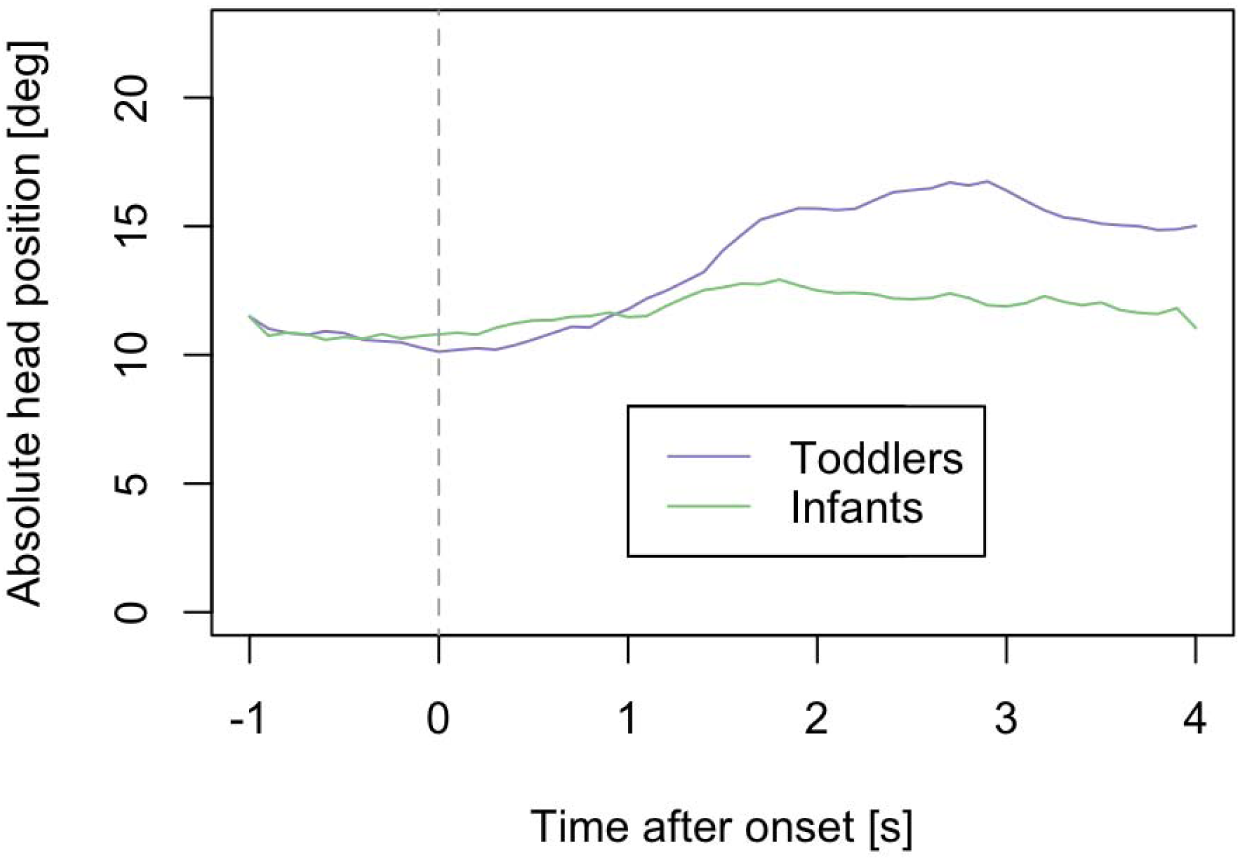
Average absolute angle of the head turns to the left or right (yaw), relative to the position of the camera, as a function of time after stimulus onset for the toddler group (purple line) and infant group (green line). Red line indicates the analysis window (2 seconds) after stimulus onset.

### 3.2. Expectation Maximization Analysis

EM was applied to four different sets of two features each: (1) Rotation of the head along the vertical axis (yaw, turning left or right) and x-axis (pitch, nodding), (2) Gaze direction in world coordinates of the left eye to the left/right and up/down, (3) Gaze angle averaged across both eyes to the left/right and up/down, (4) the first two components of a principal component analysis computed from all 55 features. Head turn results for all trials (Fig. 3) showed that most trials did not result in an observable head turn response. Most data points were around the origin, indicating lack of head turns, no gaze in any particular direction, nor any clear reaction that is reflected in the first two principal components. The gaze for both eyes is very similar to the results for the left eye only, however, not all data points are identical, indicating some gaze movements where left and right were not the same. About 90% of all cases are close to the origin, leaving 10% where the algorithm makes a decision. However, these data reflect a single frame only (2 seconds after stimulus onset), and for proof-of-concept it was more important to show that a decision was correct when the algorithm made one.

**Figure 3.**
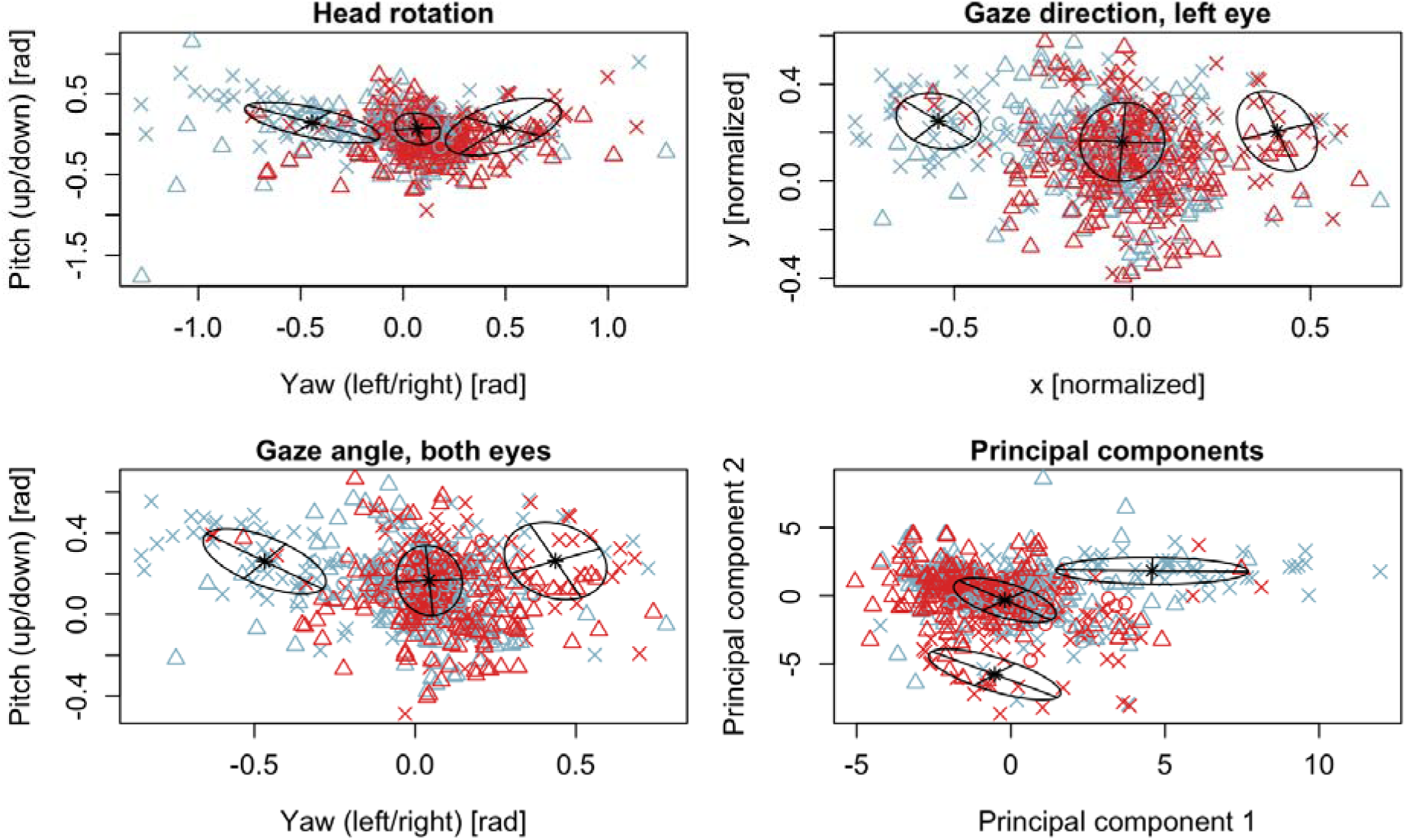
Multivariate Gaussian distributions for three clusters predicted by Expectation Maximization. Black crosses and ellipses show means and covariances for the distributions. Red symbols show reactions where a sound was presented to the right, blue symbols where a sound was presented to the left. Crosses (x) show the training data taken from the toddler group. Validation data is shown in triangles (infants) and circles (toddler).

Table 1 shows the number of predictions for each feature set, class (i.e., predicted direction) and data set (training vs. validation) separated on whether a sound was presented from the left or right speaker. This enabled us to calculate the specificity for each feature set. For the head rotation feature set, the predictive decisions were made for six out of 56 cases (ie., left or right predictive class, not centre) for the toddler validation group. Out of the six cases where a decision was made, five of these head turns were in the expected direction (i.e., sound presented from left speaker and the predictive class was left and vice versa). For the infant validation group, 13 of 24 head turns to the left followed a sound presentation from the left speaker and 31 of 52 head turns to the right followed a sound presentation from the right speaker. This results in a specificity of 58%. Using a one-sided binomial test of statistical significance (i.e., assessing the probability that at least 44 of 76 reactions are correct when guessing at 50%), it was not statistically significantly greater than a guessing rate of 50%, *p* = 0.07. The validation set of the toddlers contains too little data for meaningful statistical tests, though 5 of 6 decisions (83%) were in the expected direction.

**Table 1.** Number of predictions for each of the four feature sets and three predictive classes for the training and validation sets.

| Feature Sets | Predictive Classes | Training Set (Toddler) |  | Validation (Toddler) |  | Validation (Infant) |  | p-value |
| --- | --- | --- | --- | --- | --- | --- | --- | --- |
|  |  | Left Speaker | Right Speaker | Left Speaker | Right Speaker | Left Speaker | Right Speaker |  |
| **Head Rotation** | Left | 48 | 4 | 4 | 1 | 13 | 11 | 0.07 |
|  | Centre | 65 | 72 | 23 | 27 | 102 | 100 |  |
|  | Right | 9 | 28 | 0 | 1 | 21 | 31 |  |
| **Gaze Direction (Left Eye)** | Left | 39 | 4 | 4 | 0 | 7 | 2 | 0.006 |
|  | Centre | 78 | 84 | 23 | 29 | 127 | 131 |  |
|  | Right | 5 | 16 | 0 | 0 | 2 | 9 |  |
| **Gaze Angle (Both Eyes)** | Left | 43 | 5 | 5 | 0 | 14 | 10 | <0.001 |
|  | Centre | 67 | 78 | 22 | 28 | 116 | 114 |  |
|  | Right | 12 | 21 | 0 | 1 | 6 | 18 |  |
| **Principal Component Analysis** | Left | 45 | 4 | 5 | 0 | 15 | 10 | 0.04 |
|  | Centre | 72 | 82 | 22 | 28 | 118 | 122 |  |
|  | Right | 5 | 18 | 0 | 1 | 3 | 10 |  |

\* PCA analysis only includes the first two components

For gaze direction of the left eye, the toddler validation set showed a decision was made in 4 of the 56 cases and all 4 of those predictions were in the expected direction. For the infant group, a decision was made in 20 out of 278 cases of which 16 predictions were in the expected direction, resulting in a specificity of 80% and statistical significance, *p* = 0.006. Very similar results were found for gaze averaged across both eyes (Figure 3, bottom left panel). All 6 reactions of the toddler validation set were in the expected direction (100%) and so were 32 of the 48 reactions of the infant group, constituting a specificity of 67% and statistical significance (*p* < 0.001).

PCA captured features, or combinations of features other than head turns and gaze. Table 1 shows the performance for EM based on the first two components. For the toddler validation set, only 6 predictions were made, though all of them were in the expected direction (100%). For the infant validation group, 25 of 38 predictions were in the expected direction resulting in a specificity of 66% (*p* = 0.04).

A human observer would base a decision on large or easily observed behavioural responses like head turns and less reliably so on subtle features like eye movements.

Combining features can be a strength of more complex ML algorithms. We investigated the potential to increase sensitivity by comparing the agreement between predictions based on head movement with predictions based on the gaze averaged across both eyes. For the toddlers, in four cases a decision was made by both head movement and gaze, and they agreed in direction. There were two cases for each head movements and gaze where only one of them made a decision. Thus, there were eight cases in total where a decision was based on head movement or gaze, compared to six cases each when it was based on only one of them. For the infants, combining the 76 reactions based on head turns and 48 reactions based on gaze direction resulted only in a modest increase to 82 decisions. Of the 42 cases where both features led to a decision, 39 predictions agreed in direction. The three cases where head movement and gaze differed were correctly predicted by the gaze, and all three followed sound presentations to the left.

## 4. Discussion

The ML approach used here was naïve to the nature and direction of the stimuli (left vs. right speaker) and type of response yet was able to train an algorithm based on toddler responses to detect gaze direction and angle. In infants, head turns were detected in 27% of cases, but only 58% of those turns were toward the direction of the stimulus presentation. It was somewhat higher, 67% to 80%, when predictions were based on eye gaze, however a decision could be made in only 7% to 17% of cases. Because young infants do not generally make clear head turns, an ML approach based on a variety of facial features may be helpful during the first few months when early hearing diagnosis and initiation of rehabilitation is critically important.

The EM algorithm formed clusters without knowledge to which side the stimuli were presented. Nonetheless, three clusters always formed, one central, and one to each of the left and right, with the left and right clusters showing about equal probability, as expected. This was also true for the PCA data, which included additional information about facial movement. It therefore appears possible to obtain meaningful lateralization of the behavioural response to sound from the data alone.

For PCA analysis, a decision was made for 11% of the toddler validation group and 14% of the infant validation group. In these cases, the PCA results showed high specificity, with all six predictions correct in the toddler group and 66% for the infant group. However, this does not constitute an improvement compared to using gaze alone. PCA is a rather simple and linear approach to combine features. It remains to be seen if more complex approaches based on bigger data sets, based on 1,000 or more participants, and supervised combination of features may offer improved performance. The high specificity of only two components of the PCA, which were formed from all 55 features in an unsupervised way, is promising for such an approach. The lowest accuracy for the infant group was achieved when analysing head turns, which indicates that infants show fewer head turns than the toddlers, on whom the algorithm was trained.

A significant drawback in the present analysis is the substantial proportion of data for which no decision was made on whether the child looked to the left or to the right. This was the case for about 90% of the toddler validation data, and 73% to 93% for the infant validation data, depending on feature set. This may be partly explained by the child not reacting to the stimulus, or the reaction being finished before or starting later than 2 seconds after stimulus onset. Figure 3 shows most of the head rotation and gaze data to be focused on the computer screen or camera rather than one of the two sound field speakers. This is consistent with previous publications that showed a 32% head turn response rate for unreinforced complex noise in infants between 12 to 18 months of age (Moore et al., 1975) and only 25% of infants (6-30 months) made a reliable head turn to warble tones (Shaw & Nikolopoulos, 2004). Another possibility is that the test assistant, whose task was to keep the child looking ahead and centring the child if they were looking away, may have interfered with the response. Furthermore, a considerable number of head turns, and eye gazes were made in the direction opposite to that expected, as is evident by blue and red symbols on both sides of the figures. Theoretically, the algorithm could be improved by counting head turns in either direction, and by considering a wider window of time in which responses may occur (as opposed to a single frame 2 seconds after stimulus onset), which would allow for improved decision rates. This would require control trial (no sound) data and a comparison between control and sound trials as opposed to the left/right comparisons made in the present study. This would be consistent with traditional VRA approaches that require only detection of a sound by means of a conditioned head turn response, and do not require the head turn to be in the direction of the sound source. In other words, VRA is more sensitive when used as a detection task rather than a localization task (Widen et al., 2000). Furthermore, such techniques could be used in a VRA procedure in which a child’s head turns are reinforced which would significantly increase the turning behaviour observed and therefore improve the likelihood of appropriate response classification. Real observers can learn about the child’s response time through observation and are able to include responses that are slow but consistent. An effective AI approach would similarly be able to learn and adapt to the individual’s reaction time.

Altogether, the present analyses with simple machine learning techniques and few data demonstrate proof-of-concept and categorize the head turn direction with reasonable specificity. More data that is anonymized via preprocessing with OpenFace will allow us to employ more complex and possibly more accurate machine learning approaches, like variational autoencoders (Kingma & Welling, 2013) to replace the PCA, neural networks to capture possible non-linear interactions between the features, and a more complex AI that does not analyse a single frame but the whole-time span after stimulus onset. These approaches could improve the sensitivity of detection, enabling diagnostic and rehabilitative audiometric applications. Further improvement may be realized with data that is manually labelled with a human observer’s judgement of whether a child showed a facial reaction or not. This is likely to show higher accuracies since a detection task is easier than the present localization paradigm.

Lastly, this proof-of-concept study was conducted in infants and toddlers with normal hearing that demonstrated high completion rates on behavioural hearing assessments (60% of toddlers completed full assessments, defined as 2 speech thresholds and 4 pure-tone thresholds in each ear), which may partly explain the strong classification performance observed for the infant and toddler validation groups when a decision was made (7 to 11% toddler cases; 7 to 17% infant cases). Future work should evaluate this approach in more clinically complex populations, such as children with hearing loss and those with additional neurological or developmental conditions—groups that are often the least testable using conventional behavioural methods. Examining the clinical implications of this method, including comparisons between machine learning–based classifications and responses judged by trained clinicians, will provide valuable insight. Ongoing data collection includes full-term infants as well as a subset of infants and toddlers with hearing loss and developmental conditions, which will allow us to explore these questions in future studies.

## 5. Conclusions

1. In this proof-of-concept study, an unsupervised ML algorithm was tasked to form three meaningful clusters. The cluster representing “no decision by the algorithm” encompassed 90% of the data. When a decision was made about the direction of sound presentation (“right” and “left’ clusters), it was mostly correct for the toddlers and significantly above chance correct for the infants, despite the algorithm being trained on toddlers’ reactions.
2. Automatic VRA could be possible with unsupervised machine learning techniques and few data, as demonstrated in this study using expectation maximization. Performance level demonstrated here may be used to provide a human rater with an additional opinion.
3. The approach was applied to anonymous data, achieved by preprocessing the images to result in actions performed by the head and face rather than a description of the face. This allows us to collect and share data across clinics and apply more powerful AI techniques like deep learning to larger data sets.

## Data Availability

All data produced in the present study will be available online in a data repository at the conclusion of the study.

## Abbreviations

AI: Artificial Intelligence
BOA: Behavioural Observation Audiometry
CPA: Conditioned Play Audiometry
EM: Expectation Maximization
ML: Machine Learning
NICU: Neonatal Intensive Care Unit
OPP: Observer Psychoacoustic Procedure
PCA: Principal Component Analysis
VRA: Visual Reinforcement Audiometry

## Acknowledgements

CMB and JS are joint first author and LLH and DRM are joint senior author.

## Notes

**Funding:** Supported by NIH/NIDCD grant R01DC018734 (MPI: Hunter and Vannest), NIHR Manchester Biomedical Research Centre (NIHR BRC-20007), William Demant Foundation, and the Translational Science Award Program Grant 5UL1TR001425-04 (Center for Clinical and Translational Science Training at the University of Cincinnati). Munro is supported by an NIHR Senior Investigator award.

### Competing Interest Statement

The authors have declared no competing interest.

### Funding Statement

Supported by NIH/NIDCD grant R01DC018734 (MPI: Hunter and Vannest), NIHR Manchester Biomedical Research Centre (NIHR BRC-20007), William Demant Foundation, and the Translational Science Award Program Grant 5UL1TR001425-04 (Center for Clinical and Translational Science Training at the University of Cincinnati).

### Author Declarations

Ethics committee/IRB of Cincinnati Childrens Hospital Medical Center gave ethical approval for this work.

### Summary of Updates

We have reorganized the introduction to improve the flow and readability, expanded on the discussion of behavioral hearing assessments in infants and children, and clarified the purpose and goals of the study. We added more details to the methods/results section and expanded the discussion of future directions and limitations.

